# The Microbiome in Adult Acute Appendicitis

**DOI:** 10.1101/2022.03.11.22272248

**Authors:** Mei Sze Lee, Arielle Sulit, Frank Frizelle, Rachel Purcell

## Abstract

Acute appendicitis is a common acute surgical emergency; however, the pathogenesis of adult appendicitis remains poorly understood. The microbiome is increasingly thought to play a key role in inflammatory disease of the bowel and similarly, may play a role in appendicitis.

**Aim:** This study aimed to characterize the microbiome of adult acute appendicitis in a prospective cohort.

**Method:** We recruited 60 adults with acute appendicitis and 20 healthy controls. Rectal swabs were taken from each patient. After DNA extraction, 16S rRNA amplicon sequencing was carried out for analysis of diversity and taxonomic abundance.

**Results:** Phylogenetic sequencing of the samples indicated that there is a difference between the microbial composition of those with acute appendicitis and healthy controls, with a statistically significant decrease in alpha diversity in rectal swabs of appendicitis patients compared to healthy controls. At the genus level, we saw an increased abundance of potential pathogens, e.g. *Parvimonas* and *Acinetobacter*, and a decrease in commensal taxa such as *Faecalibacterium, Blautia* and *Lachnospiraceae* in appendicitis patients compared to healthy controls.

**Conclusion:** An imbalance in the gut microbiome may contribute to the pathogenesis of adult acute appendicitis.

## Introduction

Acute appendicitis is a common acute surgical emergency, with an estimated 17.7million cases worldwide per year ^(Wickramasinghe, 2021)^ and a lifetime risk of appendicitis of approximately 1 in 8^(Lee et al., 2010)^. The aetiology of acute appendicitis remains unknown. Direct luminal obstruction has been the conventional theory for the pathogenesis of acute appendicitis, commonly associated with faecoliths, lymphoid hyperplasia, parasites and very rarely caecal or appendiceal malignancy. Environmental factors such as seasonal alterations in temperature also affect the incidence and course of acute appendicitis ^(Wolkomir et al.)^.

The importance of the gut microbiome is increasingly being recognized in gut-related diseases, especially its role in inflammation (Duvallet et al., 2017; Glassner et al., 2020; Jackson et al., 2014; Oh et al., 2020). Several studies using bacterial cultures of paediatric populations have highlighted the association of various microbiota in the pathogenesis of acute appendicitis ^(Elhag et al.; Hattori et al., 2019; Swidsinski et al.)^. The advancement of high throughput sequencing of the human microbiome has led to a growth in knowledge of the microbiome and the role of dysbiosis in disease. Despite this, there has been limited research undertaken on adult appendicitis. The aim of this study is to characterize the microbiome of adult acute appendicitis using 16sRNA gene sequencing in a prospective cohort.

## Methods

### Participants

Patients who presented with acute appendicitis and subsequently underwent laparoscopic appendectomy, between March and September 2020, were recruited prospectively at Christchurch Hospital, Canterbury, New Zealand. Patients had a rectal swab taken pre-operatively. Demographics and relevant clinical data were collected from electronic hospital health system. Participants were 18 years and older, with acute appendicitis confirmed on histology for inclusion in the study. Patients were excluded if they had antibiotic for more than 24 hours prior to surgery or in the previous six weeks. Appendicectomies performed in conjunction with concurrent colorectal or gynaecological disease were also excluded. Healthy volunteers had rectal swabs taken by a single clinician. This study had approval from the University of Otago Human Ethics Committee (H20/045). All participants provided written informed consent upon enrollment.

### DNA extraction and 16S rRNA sequencing

All swabs were stored in DNA/RNA lysis tubes (Zymo Research, California, USA) to maintain optimum stability of the DNA. DNA was extracted from samples using ZymoBIOMICS micro-DNA Kits (Zymo Research), according to the manufacturer’s instructions. In brief, samples homogenized using ZR BashingBead followed by centrifugation at >10,000 rpm for a minute. Supernatant was added to DNA Binding buffer and transferred Zymo-Spin IC-Z Column in a collection tube and centrifuged at maximum for one minute. The column was washed several times and DNA eluted in 20µl DNase/RNase-free water. Finally, DNA concentration and 260/280 nm absorbance were measured using the NanoDrop 2000 spectrophotometer (Thermo Fisher Scientific, Massachusetts, USA). DNA was stored at −20C prior to 16S analysis.

### Microbiome Analysis

#### Quality Control

DADA2 (v1.18.0) was used to filter, trim, join sequencing reads, and remove chimeras to obtain amplicon sequence variants^(Callahan et al., 2016)^. The SILVA database (v132) was utilised to assign taxonomy to the amplicon sequence variants^(Quast et al., 2013)^. The taxonomy, sample metadata, and sequences were combined into a phyloseq object for subsequent analysis^(McMurdie & Holmes, 2013)^. Samples with less than 1000 reads after DADA2 quality control were dropped.

#### Phylum-level Differences

Taxa identified were agglomerated to the Phylum level and percent abundances were obtained per samples. Wilcoxon tests were used to compare Phylum differences between groups. A p-value of < 0.05 is considered significant.

#### Diversity Analysis

We rarefied samples to the smallest library size. We used phyloseq’s estimate richness function for Observed (richness) and Shannon (richness and evenness) measures, and compared differences of the alpha diversity measures by Wilcoxon tests. A p-value of < 0.05 is considered significant. We used principal coordinate analysis (PCoA) to test for overall differences in the microbiome between groups, using UniFrac distances between samples.

#### Differential abundance

DESeq2 was used to identify differentially abundant genera between healthy and appendicitis groups. Only genera that had 10% abundance in 10% of samples from each group were used. We considered a fold-change between comparisons significant if it has adjusted p-value of < 0.05.

## Results

A total of 60 patients with acute appendicitis and 20 healthy controls were recruited for the study. Four patients did not have sufficient samples materials for analysis. The median age was 35.5 years old in appendicitis group and 32.5 years old in the control group.

In the appendicitis group, participants were predominantly male with NZ European ethnicity. There were 31 patients with simple appendicitis and 25 patients (44.6%) with complicated appendicitis (perforated and/or gangrenous). Patients were older and had slightly higher inflammatory markers in the complicated appendicitis subgroup. **(Table 1)**

**Table 1:**
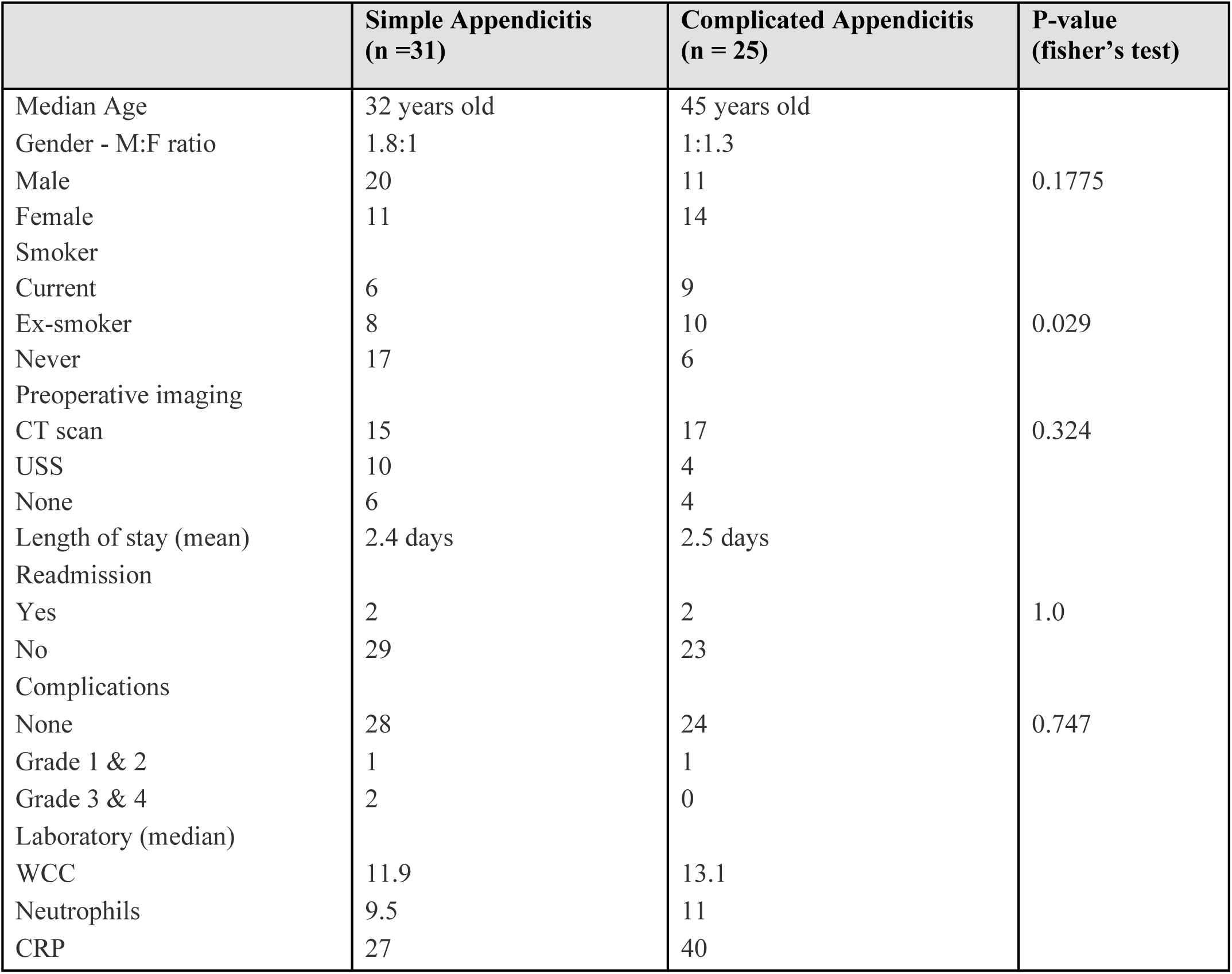
Demographics of the appendicitis group.

### Comparison of the microbiome of acute appendicitis and healthy group

Bioinformatic analysis showed a statistically significant difference in the alpha diversity between the groups (p < 0.0001), with samples from healthy controls having higher alpha diversity. Unweighted UniFrac analysis showed clustering of samples by appendicitis or health controls **(Figure 3A)**. Wilcoxon tests on phyla between healthy and appendicitis sample showed a significant difference of *Firmicutes, Actinobacteria* and *Proteobacteria* phyla percent abundance. Further classification at the genus level show loss of beneficial gut commensals, including *Blautia, Lachnospiraceae, Faecalibacterium and Ruminococcus* in swabs from appendicitis patients, compared to healthy controls **(Figure 3B)**.

**Figure 3A:**
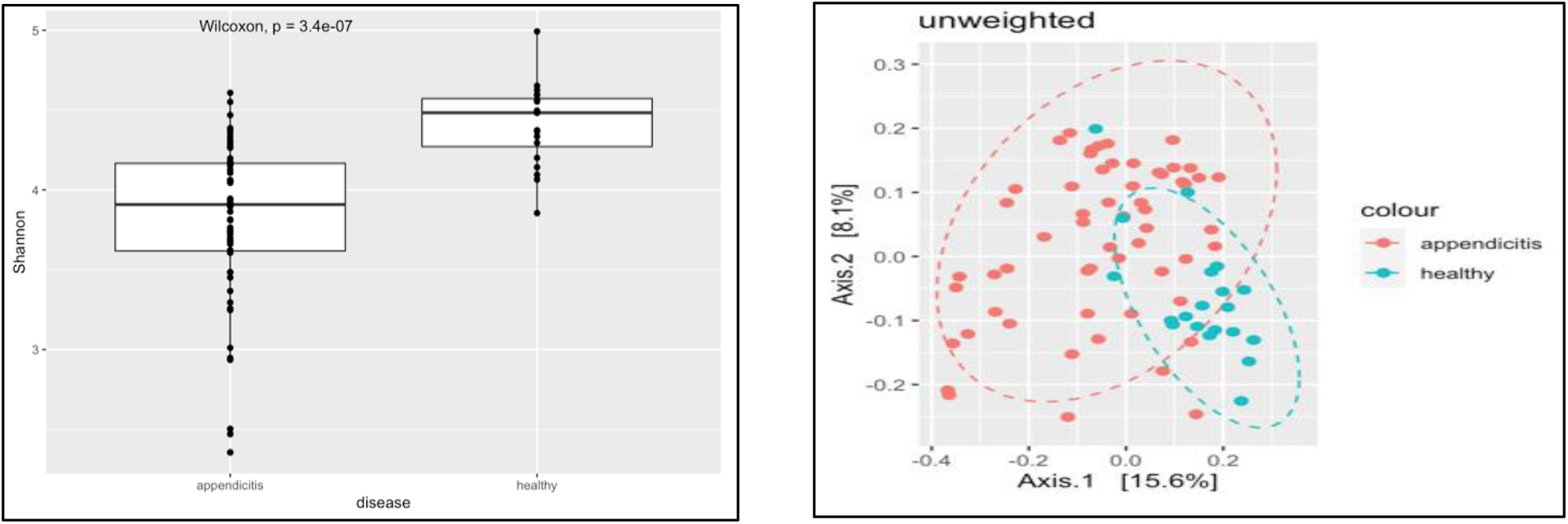
Alpha diversity and PCoA of rectal swabs between appendicitis and healthy controls.

**Figure 3B:**
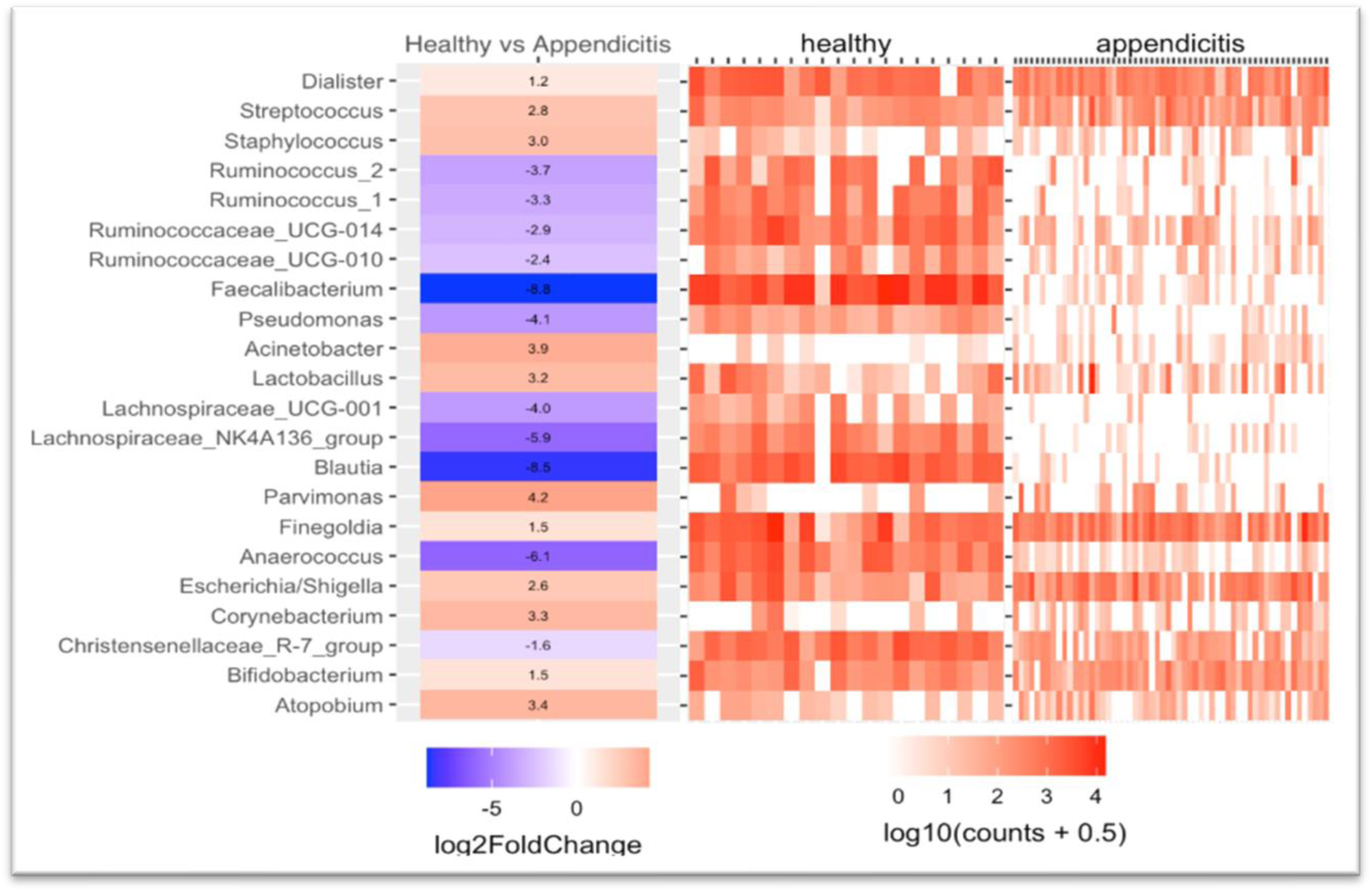
Comparison between appendicitis and healthy samples at genus level. A positive log fold change means the genus more abundant in appendicitis and a negative log fold means the genus is more abundant in healthy samples.

### Comparison of the microbiome between simple and complicated appendicitis

Shannon diversity analysis showed no significant differences in alpha diversities between simple and complicated appendicitis in rectal samples (p = 0.98) **(Figure 4A)**. Phylum-level abundance were similar for both simple and complicated acute appendicitis. Further classification at the genus level showed no difference between the two groups.

**Figure 4A:**
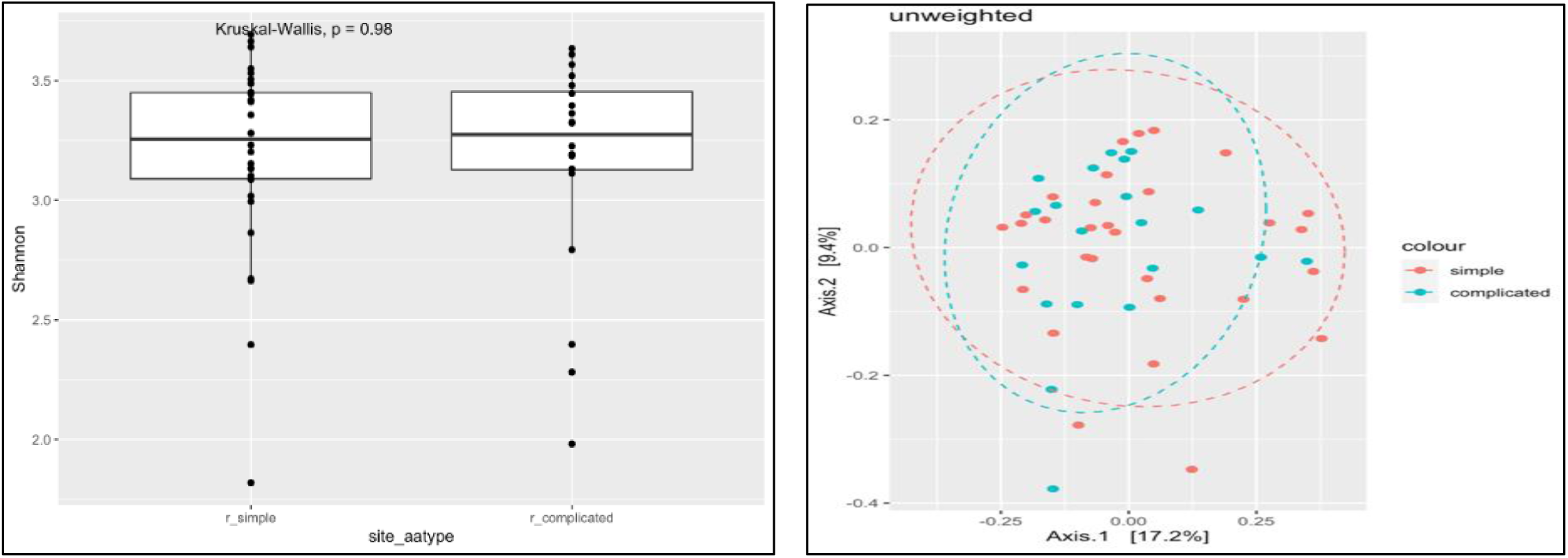
Alpha diversity and PCoA of rectal swabs comparing simple and complicated appendicitis.

**Figure 4B:**
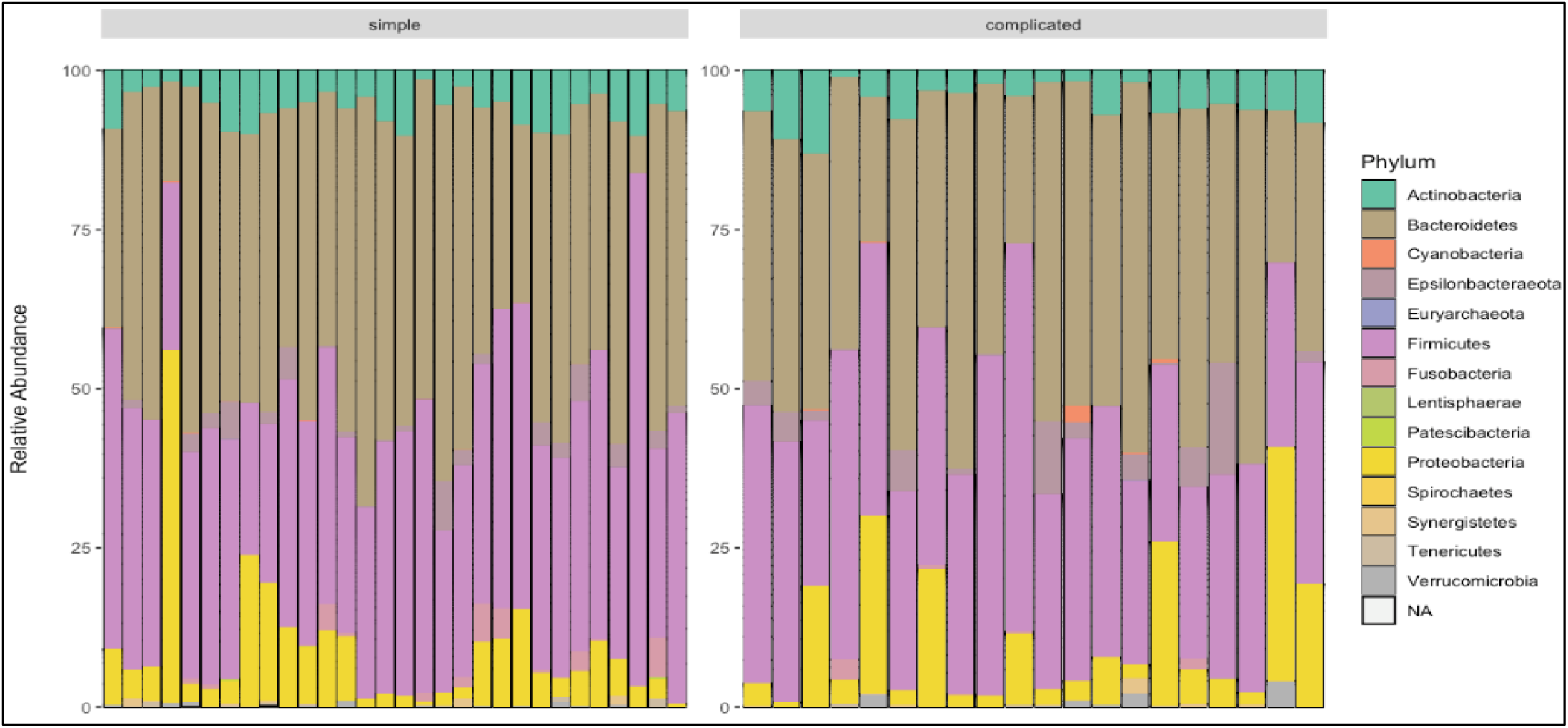
Phyla distribution between simple and complicated appendicitis.

## Discussion

In this study, we found that there is a difference between the microbial composition as assessed by rectal swabs of those with acute appendicitis and healthy controls. Metagenomics sequencing analysis demonstrates a reduction of commensal bacteria in people with appendicitis. There were no significant microbiome differences found between patients with simple and complicated appendicitis.

Despite the frequency of acute appendicitis, we know very little about its aetiology. Microbiological causes have been suggested in various studies. Guinane et al^(Guinane et al., 2013)^ first provided insight into the microbial composition of the appendix in a study of seven patients, aged 5 to 25 years old, using 16S-rRNA sequencing. Analysis of faecal and tissue samples revealed that appendices contained a high bacterial biomass and were postulated to be a reservoir for commensal bacterial. Across the seven samples, 15 different phyla were detected, dominated by five major phyla – *Firmicutes, Proteobacteria, Bacteroidetes, Actinobacteria* and *Fusobacteria*. In our study of an adult cohort, we found similar dominant phyla with the exception of *Fusobacteria*.

Several other paediatric studies of appendicitis suggest that *Fusobacterium* is associated with acute appendicitis(Salo et al., 2017; Schulin et al., 2017; Swidsinski et al., 2012; Zhong et al., 2014). Salo et.al. noticed an increased abundance of *Fusobacterium* and a decreased in *Bacteroides* in complicated appendicitis compared to uncomplicated appendicitis^(Salo et al., 2017)^. A further study of intraluminal samples by Schulin et al. reported that *Fusobacterium necrophorum* was mainly found in catarrhal appendicitis, *Pseudomonas endodontalis* in phlegmonous appendicitis and *F*.*nucleatum* in gangrenous appendicitis^(Schulin et al., 2017)^. In contrast in our study, *Fusobacterium* was not one of the predominant phyla in the adult appendicitis and healthy controls, which may reflect very different methodologies.

A growth in human microbiome research has led to better understanding of disease associated microbiome change. A study on the aetiology of appendicitis in 277 children showed that *Escherichia coli* was the predominant bacteria followed by *Streptococci*^(Richardsen et al., 2016)^. In addition, Zhong et al. performed the 16S ribosomal gene sequence analysis to profile the microbiota of the luminal fluid of the appendix in children with appendicitis and normal appendices, and found an increased relative abundance of the pathogen containing genera, *Fusobacterium* and *Prevotella* with a corresponding depletion of *Bacteroides* (11.4% vs 52.9%) ^(Zhong et al., 2014)^. In our study, *Actinobacteria* and *Proteobacteria* phyla are increased in appendicitis patients with a concomitant increase in the pathogen containing *Parvimonas, Acinetobacter, E. coli, Corneybacterium* and *Streptococci* genera. The apparent differences in microbiome changes between paediatric and adult population might be due to different methodologies in the various studies or other factors (i.e dietary or lifestyle factors) which may influence the dysbiosis in acute appendicitis.

Analysis of differential abundance at the genus level in our study also revealed a loss of commensal bacteria in appendicitis samples compared to healthy samples. These included *Blautia, Faecalibacterium and Lachnospiracaea*, genera that contain short-chain fatty acids (SCFA) producing bacteria. SCFAs are important metabolites that are an energy source for epithelial cells and also contribute to gut barrier function^(Loh & Blaut, 2012;^ Parada Venegas et al., 2019) and play an anti-inflammatory role in the gut (Blaak et al., 2020; Machiels et al., 2014; Segain JP, 2000). Our observed reduction of microbial diversity has been described in other inflammatory conditions such as inflammatory bowel disease and acute appendicitis^(Loh & Blaut, 2012)^. Ott et. al performed a 16S rDNA study of mucosa associated colonic microflora and found a reduction of microbial diversity in both Crohn’s disease and Ulcerative colitis^(Ott, 2004)^. This demonstrates that gut inflammation has a strong association with changes in gut microbiome.

This is one of the few published studies on the microbiome of acute appendicitis in adult population. We have used rectal swabs instead of faecal sampling a methodology we have previously verified ^(Budding et al., 2014)^. The methodology of this study does not however allow us to determine whether gut microbiome dysbiosis was a cause of appendicitis or just secondary to the pathology. Despite the relatively large cohort size in this study, future large-scale studies are necessary to validate the findings presented here and to establish bacterial signatures associated with acute appendicitis.

## Conclusion

In conclusion, this study has added knowledge to the current literature on microbiome studies of acute appendicitis in adult population. We have found a reduction in diversity and loss of commensals in the microbiome of those with acute appendicitis. The loss of commensals bacteria may play a role in the cascade leading to the development of acute appendicitis.

## Data Availability

Research Transparency and Reproducibility: Protocols, materials and relevant data are available for readers and can be access by contacting corresponding authors.

## Acknowledgements

The authors would like to thank University of Otago administrative staffs.

## Author contributions

**Conceptualisation:** F.F **Data curation:** M.S.L **Formal analysis:** A.S, R.P **Investigations:** A.S, R.P

**Methodology:** M.S.L, R.P, A.S **Project administration:** F.F **Resources:** M.S.L

**Software:** A.S

**Visualisation:** A.S, M.S. L

**Writing – original draft:** M.S.L **Writing – review & editing:** R.P, F.F **Supervision:** R.P, F.F

## Financial support

No external funders.

## Disclosure statement

The authors have no known conflicts of interest to report.

## Research Transparency and Reproducibility

Protocols, materials and relevant data are available for readers and can be access by contacting corresponding authors.

## Notes

### Competing Interest Statement

The authors have declared no competing interest.

### Funding Statement

Financial support: This study did not receive any external funders.

### Author Declarations

This study had approval from the University of Otago Human Ethics Committee (H20/045).

## References

Blaak, E. E., Canfora, E. E., Theis, S., Frost, G., Groen, A. K., Mithieux, G., Nauta, A., Scott, K., Stahl, B., van Harsselaar, J., van Tol, R., Vaughan, E. E., & Verbeke, K. (2020). Short chain fatty acids in human gut and metabolic health. Benef Microbes, 11(5), 411–455. https://doi.org/10.3920/BM2020.0057

Budding, A. E., Grasman, M. E., Eck, A., Bogaards, J. A., Vandenbroucke-Grauls, C. M., van Bodegraven, A. A., & Savelkoul, P. H. (2014). Rectal swabs for analysis of the intestinal microbiota. PLoS ONE [Electronic Resource], 9(7), e101344. https://doi.org/10.1371/journal.pone.0101344

Callahan, B. J., McMurdie, P. J., Rosen, M. J., Han, A. W., Johnson, A. J., & Holmes, S. P. (2016). DADA2: High-resolution sample inference from Illumina amplicon data. Nat Methods, 13(7), 581–583. https://doi.org/10.1038/nmeth.3869

Duvallet, C., Gibbons, S. M., Gurry, T., Irizarry, R. A., & Alm, E. J. (2017). Meta-analysis of gut microbiome studies identifies disease-specific and shared responses. Nature Communications, 8(1784), 1–10. https://doi.org/10.1038/s41467-017-01973-8

Glassner, K. L., Abraham, B. P., & Quigley, E. M. M. (2020). The microbiome and inflammatory bowel disease. Journal of Allergy and Clinical Immunology, 145(1), 16–27. https://doi.org/10.1016/j.jaci.2019.11.003

Guinane, C. M., Tadrous, A., Fouhy, F., Ryan, C. A., Dempsey, E. M., Murphy, B., Andrews, E., Cotter, P. D., Stanton, C., & Ross, R. P. (2013). Microbial composition of human appendices from patients following appendectomy. mBio, 4(1), 1–6. https://doi.org/10.1128/mBio.00366-12

Hattori, T., Yuasa, N., Ikegami, S., Nishiyama, H., Takeuchi, E., Miyake, H., Kuno, R., Miyata, K., Fujino, M., & Minami, M. (2019). Culture-based bacterial evaluation of the appendix lumen in patients with and without acute appendicitis. Journal of Infection & Chemotherapy, 25(9), 708–713. https://dx.doi.org/10.1016/j.jiac.2019.03.021

Jackson, H. T., Mongodin, E. F., Davenport, K. P., Fraser, C. M., Sandler, A. D., & Zeichner, S. L. (2014). Culture-independent evaluation of the appendix and rectum microbiomes in children with and without appendicitis. PLoS ONE [Electronic Resource], 9(4), e95414. https://doi.org/10.1371/journal.pone.0095414

Lee, J. H., Park, Y. S., & Choi, J. S. (2010). The epidemiology of appendicitis and appendectomy in South Korea: national registry data. J Epidemiol, 20(2), 97–105. https://doi.org/10.2188/jea.je20090011

Loh, G., & Blaut, M. (2012). Role of commensal gut bacteria in inflammatory bowel diseases. Gut Microbes, 3(6), 544–555. https://doi.org/10.4161/gmic.22156

Machiels, K., Joossens, M., Sabino, J., De Preter, V., Arijs, I., Eeckhaut, V., Ballet, V., Claes, K., Van Immerseel, F., Verbeke, K., Ferrante, M., Verhaegen, J., Rutgeerts, P., & Vermeire, S. (2014). A decrease of the butyrate-producing species Roseburia hominis and Faecalibacterium prausnitzii defines dysbiosis in patients with ulcerative colitis. Gut, 63(8), 1275–1283. https://doi.org/10.1136/gutjnl-2013-304833

McMurdie, P. J., & Holmes, S. (2013). phyloseq: an R package for reproducible interactive analysis and graphics of microbiome census data. PLoS ONE [Electronic Resource], 8(4), e61217. https://doi.org/10.1371/journal.pone.0061217

Oh, S. J., Pimentel, M., Leite, G. G. S., Celly, S., Villanueva-Millan, M. J., Lacsina, I., Chuang, B., Parodi, G., Morales, W., Weitsman, S., Singer-Englar, T., Barlow, G. M., Zhai, J., Pichestshote, N., Rezaie, A., Mathur, R., & Pimentel, M. (2020). Acute appendicitis is associated with appendiceal microbiome changes including elevated Campylobacter jejuni levels. BMJ Open Gastroenterol, 7(1), 1–10. https://doi.org/10.1136/bmjgast-2020-000412

Ott, S. J. (2004). Reduction in diversity of the colonic mucosa associated bacterial microflora in patients with active inflammatory bowel disease. Gut, 53(5), 685–693. https://doi.org/10.1136/gut.2003.025403

Parada Venegas, D., De la Fuente, M. K., Landskron, G., Gonzalez, M. J., Quera, R., Dijkstra, G., Harmsen, H. J. M., Faber, K. N., & Hermoso, M. A. (2019). Short Chain Fatty Acids (SCFAs)-Mediated Gut Epithelial and Immune Regulation and Its Relevance for Inflammatory Bowel Diseases. Frontiers in Immunology, 10(277), 1–16. https://doi.org/10.3389/fimmu.2019.00277

Quast, C., Pruesse, E., Yilmaz, P., Gerken, J., Schweer, T., Yarza, P., Peplies, J., & Glockner, F. O. (2013). The SILVA ribosomal RNA gene database project: improved data processing and web-based tools. Nucleic Acids Res, 41(Database issue), D590–596. https://doi.org/10.1093/nar/gks1219

Richardsen, I., Schob, D. S., Ulmer, T. F., Steinau, G., Neumann, U. P., Klink, C. D., & Lambertz, A. (2016). Etiology of Appendicitis in Children: The Role of Bacterial and Viral Pathogens. J Invest Surg, 29(2), 74–79. https://doi.org/10.3109/08941939.2015.1065300

Salo, M., Marungruang, N., Roth, B., Sundberg, T., Stenstrom, P., Arnbjornsson, E., Fak, F., & Ohlsson, B. (2017). Evaluation of the microbiome in children’s appendicitis. Int J Colorectal Dis, 32(1), 19–28. https://doi.org/10.1007/s00384-016-2639-x

Schulin, S., Schlichting, N., Blod, C., Opitz, S., Suttkus, A., Stingu, C. S., Barry, K., Lacher, M., Buhligen, U., & Mayer, S. (2017). The intra-and extraluminal appendiceal microbiome in pediatric patients: A comparative study. Medicine (Baltimore), 96(52), e9518. https://doi.org/10.1097/MD.0000000000009518

Segain JP, d. l. B. D., Bourreille A, Leray V, Gervois N et. al. (2000). Butyrate inhibits inflammatory responses through NFkB inhibition: implications for Crohn’s disease. Gut Journal, 47, 397–403.

Swidsinski, A., Dorffel, Y., Loening-Baucke, V., Theissig, F., Ruckert, J. C., Ismail, M., Rau, W. A., Gaschler, D., Weizenegger, M., Kuhn, S., Schilling, J., & Dorffel, W. V. Acute appendicitis is characterised by local invasion with Fusobacterium nucleatum/necrophorum. Gut, 60(1), 34–40. https://ovidsp.ovid.com/ovidweb.cgi?T=JS&CSC=Y&NEWS=N&PAGE=fulltext&D=med8&AN=19926616

Swidsinski, A., Loening-Baucke, V., Biche-ool, S., Guo, Y., Dörffel, Y., Tertychnyy, A., Stonogin, S., & Sun, N.-D. (2012). Mucosal invasion by fusobacteria is a common feature of acute appendicitis in Germany, Russia, and China. Saudi Journal of Gastroenterology, 18(1), 55–58. https://doi.org/10.4103/1319-3767.91734

Wickramasinghe, D. P. X., C. Samarasekera, D. N. (2021). The Worldwide Epidemiology of Acute Appendicitis: An Analysis of the Global Health Data Exchange Dataset. World J Surg, 45(7), 1999–2008. https://doi.org/10.1007/s00268-021-06077-5

Wolkomir, A., Kornak, P., Elsakr, M., & McGovern, P. Seasonal variation of acute appendicitis: a 56-year study. Southern Medical Journal, 80(8), 958–960. https://ovidsp.ovid.com/ovidweb.cgi?T=JS&CSC=Y&NEWS=N&PAGE=fulltext&D=med2&AN=3616723

Zhong, D., Brower-Sinning, R., Firek, B., & Morowitz, M. J. (2014). Acute appendicitis in children is associated with an abundance of bacteria from the phylum Fusobacteria. J Pediatr Surg, 49(3), 441–446. https://doi.org/10.1016/j.jpedsurg.2013.06.026

